# SARS-CoV-2 mRNA vaccination fails to elicit humoral and cellular immune responses in multiple sclerosis patients receiving fingolimod

**DOI:** 10.1101/2022.02.06.22270550

**Authors:** Lil Meyer-Arndt, Julian Braun, Florent Fauchere, Kanika Vanshylla, Lucie Loyal, Larissa Henze, Beate Kruse, Manuela Dingeldey, Karsten Jürchott, Maike Mangold, Ardit Maraj, Andre Braginets, Chotima Böttcher, Andreas Nitsche, Kathrin de la Rosa, Christoph Ratswohl, Birgit Sawitzki, Pavlo Holenya, Ulf Reimer, Leif E. Sander, Florian Klein, Friedemann Paul, Judith Bellmann-Strobl, Andreas Thiel, Claudia Giesecke-Thiel

## Abstract

SARS-CoV-2 mRNA vaccination of healthy individuals is highly immunogenic and protective against severe COVID-19. However, there are limited data on how disease-modifying therapies (DMTs) alter SARS-CoV-2 mRNA vaccine immunogenicity in patients with autoimmune diseases. Here we investigated the induction and stability of vaccine-specific antibodies, B cells, and T cells in multiple sclerosis (MS) patients on different DMTs in a prospective cohort study up to 6 months after homologous prime-boost mRNA vaccination. We analysed 103 MS patients of which 86 received anti-CD20-based B cell depletion (aCD20-BCD), fingolimod, interferon-β, dimethyl fumarate, glatiramer acetate, teriflunomide or natalizumab, and compared them to 17 untreated MS patients. In contrast to all other DMTs and untreated patients, treatment with aCD20-BCD or fingolimod significantly reduced anti-S1 IgG, serum neutralizing activity, and RBD- and S2-specific B cells. MS patients receiving fingolimod additionally lacked S1- and S2-reactive CD4^+^ T cell responses. The duration of fingolimod treatment, rather than peripheral blood B and T cell counts prior to vaccination, determined whether patients successfully developed humoral immune responses. Fingolimod blocks the ability of immune cells to recirculate and migrate within secondary lymphoid organs demonstrating that functional immune responses require not only immune cells themselves but also access of these cells to the site of inoculation and their unimpeded movement. The absence of humoral and T cell responses in fingolimod-treated MS patients suggests that these patients are at risk for severe SARS-CoV-2 infections despite vaccination, which is highly relevant for clinical decision-making and adapted protective measures, particularly in light of additional recently approved S1P receptor antagonists for MS treatment.

## Introduction

Coronavirus disease 2019 (COVID-19) is caused by severe acute respiratory syndrome coronavirus type 2 (SARS-CoV-2) and presents with a wide variety of symptoms ranging from asymptomatic to severe and even fatal disease courses. In addition to age and lifestyle-related diseases, other major risk factors for severe COVID-19 disease courses include immunosuppression due to chronic diseases or immunosuppressive therapies^1,2^. For patients with the autoimmune disease multiple sclerosis (MS), studies have produced mixed results regarding patient susceptibility and severity of COVID-19 mainly due to differences between the treatment^3,4,5,6^. In particular, aCD20-BCD therapies with rituximab and ocrelizumab and the sphingosine-1-phosphate (S1P) receptor functional antagonist fingolimod increased the risk of infection, hospitalization, and fatality^6,7^. Fortunately, vaccines against SARS-CoV-2 became available within a year after the new virus strain emerged and two mRNA vaccines, BNT162b2 (Pfizer-BioNTech) and mRNA-1273 (Moderna), demonstrated strong immunogenicity, efficacy, and safety in their corresponding clinical trials receiving approval at the end of 2020^8,9^. Due to their increased risk for severe COVID-19, MS patients were prioritized for vaccination^10^, however, it was expected that the strongly immunomodulatory MS treatments compromise the immunogenicity of the vaccine and alter protection^11,12^. Initial results demonstrated that MS patients treated with aCD20-BCD showed lower to absent SARS-CoV-2 spike glycoprotein-specific antibody and B cell responses. Functional T cell responses were maintained^13,14^ but with partial selective defects in antigen-specific follicular helper T cells^13^. Impaired humoral responses were also noted for patients on fingolimod treatment^15^. Despite these selected reports of reduced immunogenicity, the effects of the broad spectrum of MS treatment regimens on the humoral and cellular immune responses elicited by COVID-19 vaccine have not yet been studied in detail^16^.

In this study, we aimed to assess the clinical safety of SARS-CoV-2 vaccination as well as vaccine-induced humoral and cellular immune responses in MS patients treated with aCD20-BCD, fingolimod, interferon(IFN)-β, glatiramer acetate (GA), dimethyl fumarate (DMF), teriflunomide (TFN), or the α4-integrin monoclonal antibody natalizumab (NTZ) compared to those without any immunomodulatory treatment. Due to the high prevalence of MS and the tendency to start immunomodulatory treatment early, investigation on immunogenicity in this patient group becomes increasingly relevant for clinical practice and vaccine development.

## Methods

### Participants and ethics

The study was approved by the ethics committee of the Charité – Universitätsmedizin Berlin (EA/152/20) and was conducted in accordance with the World Medical Association’s Declaration of Helsinki of 1964 and its later amendments. Written informed consent was obtained from all participants. Participants were recruited at the MS outpatient clinic of the Charité – Universitätsmedizin Berlin between January and October 2021. Inclusion criteria were: (1) MS diagnosis according to the McDonald criteria of 2017, (2) stable disease for at least three months (no clinical progress or new symptoms, no disease activity on brain MRI), (3) continuous immunomodulatory treatment or no treatment for at least three months, (4) immunomodulatory monotherapy (if treated) and (5) no medical contraindications against SARS-CoV-2 vaccination. Out of 113 patients included, three with signs of previous SARS-CoV-2 infection (previous SARS-CoV-2 infection verified by PCR or positive anti-S1 IgG levels at baseline) and seven vaccinated with a vector-based SARS-CoV-2 vaccine were excluded from analyses resulting in a total of 103 participants (**Fig. 1a**). Blood samples, nasopharyngeal swabs and a medical history were provided at up to four time points: before primary vaccination (BL) and ∼1 month (1m; before second vaccination), ∼3 months (3m), and ∼6 months (6m) after primary vaccination (**Fig. 1b**). For 15 patients, only follow-up samples were available. All participants received a homologous prime-boost COVID-19 vaccination schedule with an mRNA vaccine (BNT162b2 (BioNTech/Pfizer) or mRNA-1273 (Moderna)). The following patients were excluded from further analyses due to dropout, SARS-CoV-2 infection or early additional (booster) vaccinations: aCD20-BCD (n=2 after 1m, n=4 after 3m), fingolimod (n=4 after 6m), IFNβ (n=2 after 3m), NTZ (n=2 after 3m). For details on assessment of vaccine reactogenicity and severe adverse events see Supplemental Methods.

**Fig. 1:**
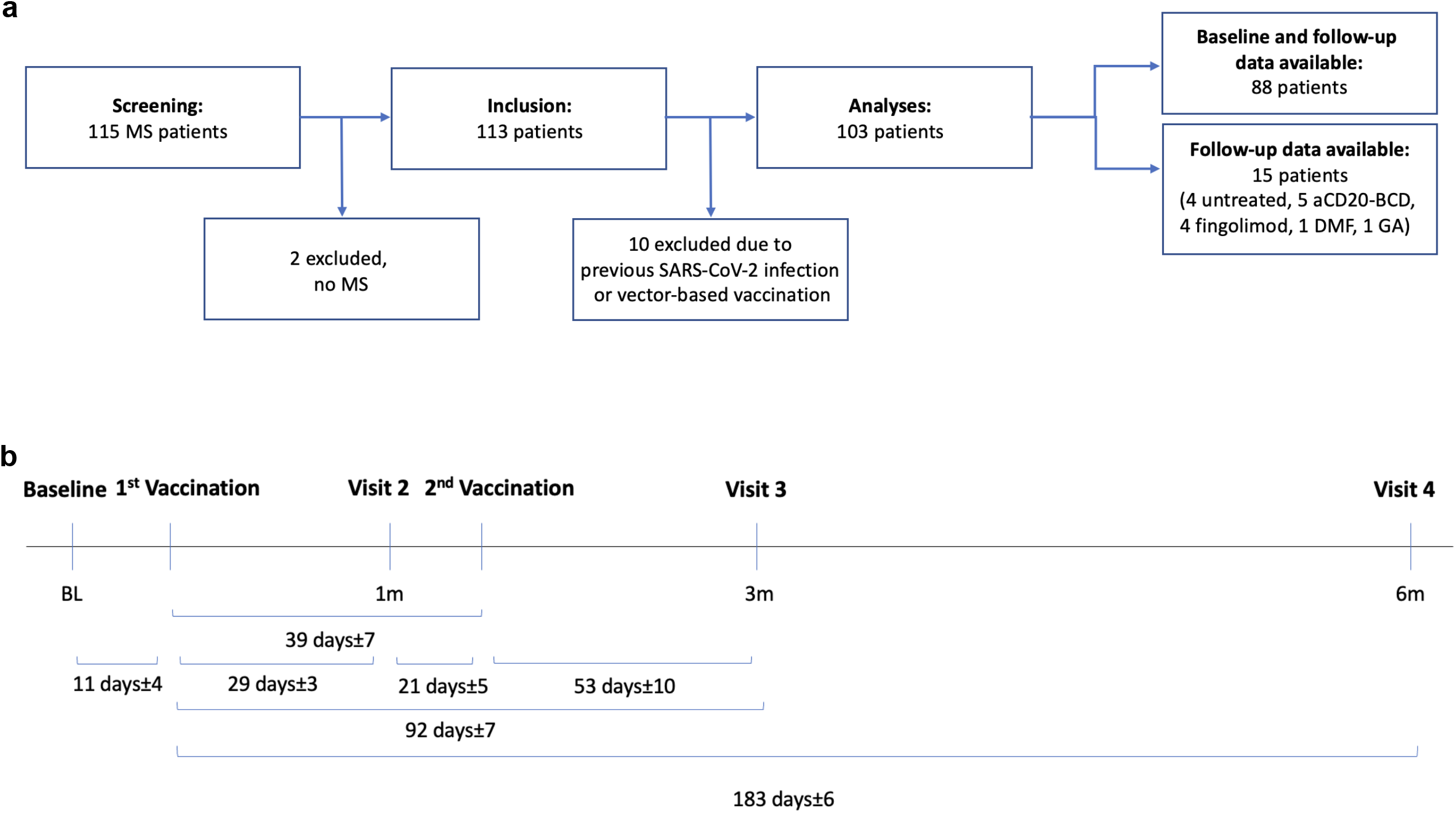
Study design and participant flow chart. **a**, timeline depicting study visits including clinical assessment and sample collection in relation to vaccinations. Time intervals reported as mean ± range across all treatment groups as no significant differences between groups were detected. **b**, participant chart showing available data for screening, data collection and analyses steps.

### SARS-CoV-2 RT-qPCR

RT-qPCR analysis of nasopharyngeal swabs was performed as previously described^17^.

### Blood sampling, serum preparation and PBMC isolation

Whole blood was collected in lithium heparin tubes for peripheral blood mononuclear cell (PBMC) isolation and in SST™II advance (all Vacutainer®, BD) tubes for serology. SST™II advance tubes were centrifuged at 1000xg for 10 min and serum supernatant aliquots were frozen at −80°C until further use. PBMCs were isolated by gradient density centrifugation using Bicoll (Bio&SELL) and Leucosep tubes (Greiner) according to the manufacturer’s instructions and immediately used for T cell stimulations and flow cytometry assays.

### Anti-SARS-CoV-2 S1 enzyme-linked immunosorbent assay (ELISA)

Anti-SARS-CoV-2 S1 IgG testing was performed using a commercially available ELISA kit (Euroimmun).

### SARS-CoV-2 pseudovirus neutralization assay

Serum neutralization was measured using a lentivirus-based pseudovirus neutralization assay as described before^18^ and in Supplemental Methods.

### SARS-CoV-2 spike epitope-specific peptide microarray

Serum IgG reactivity profiling was performed by JPT Peptide Technologies with a peptide microarray (JPT Peptide Technologies GmbH) containing 562 peptides derived from a peptide scan (15mers with 11 aa overlap) through spike glycoprotein (Swiss-Prot ID: P0DTC2) of the Wuhan type SARS-CoV-2. For a detailed method description see Supplemental Methods.

### *Ex vivo* T cell stimulations

Freshly isolated PBMC were cultivated at 5×10^6^ PBMC in RPMI 1640 medium (Gibco) supplemented with 10% heat inactivated AB serum (Pan Biotech), 100 U/ml penicillin (Biochrom), 0.1 mg/ml streptomycin (Biochrom). Stimulations with peptide pools covering the SARS-CoV-2 spike S1 subunit (S-I) and the spike S2 subunit (S-II) were conducted as described before^19,20^ and in Supplemental Methods.

### Protein labeling for RBD- and S2-specific B cells

Recombinant SARS-CoV-2 spike-S2 (Miltenyi) was covalently labelled with Pacific Blue (Antibody Labeling Kit, Invitrogen) and AlexaFluor488 (Antibody Labeling Kit, Invitrogen), respectively, and recombinant SARS-CoV-2 spike-RBD (Miltenyi) was labelled with AlexaFluor647 (Antibody Labeling Kit, Invitrogen) following the manufacturer’s instructions. For labelling of SARS-CoV-2 spike-RBD with PE-Vio615, biotinylated SARS-CoV-2 spike-RBD (Miltenyi) was incubated for 15 minutes at room temperature with Streptavidin-PE-Vio615 (Miltentyi) at 4:1 molar ratio each time before staining.

### B cell and T cell flow cytometry

Surface staining was performed for 15 min in the presence of 1mg/ml beriglobin (CSL Behring) with the following fluorochrome-conjugated antibodies titrated to their optimal concentrations. For RBD- and S2-specific B cell analysis: CD20-Viogreen (LT20, Miltenyi), CD14 BV570 (M5E2, Biolegend), CD38 BV605 (HB7, Biolegend), CD27 PE (O323, Biolegend), IgD Percp-Cy5.5 (IA6-2, Biolegend), CD19 PE-Cy7 (SJ25C1, Biolegend) and CD3 APC-Cy7 (UCHT1, Biolegend) and the labelled proteins (S2 PacBlue (0.25 µg), S2 AlexaFluor488 (0.25 µg), RBD Biotin/Streptavidin PE-Vio615 (0.15 µg) and RBD AlexaFluor647 (0.15 µg)). During the last 10 min of incubation, Zombie Yellow fixable viability staining (Biolegend) was added. After staining, the cells were washed once in PBS/BSA, centrifuged and resuspend in PBS/BSA/2mM EDTA. For analyses of S-I- and S-II-specific T cells and peripheral blood B and T cell subsets, antibody staining was performed as described before^19,20^ and in Supplemental Methods. All samples were measured on a MACSQuant® Analyzer 16 (Miltenyi). Instrument performance was monitored using Rainbow Calibration Particles (BD).

### Data collection and statistical analysis

Study data were collected and managed using REDCap electronic data capture tools hosted at the Charité. Flow cytometry data were analysed using FlowJo 10 (BD). Prism 9 (GraphPad) was used for data plotting and statistical analyses: Fisher’s exact test for seroconversion rate comparison; Kruskal-Wallis test followed by Dunn’s multiple comparisons test for group comparisons; Spearman correlation for correlation testing. Correlation analysis was performed in R (R Core Team (2021). R: A language and environment for statistical computing. R Foundation for Statistical Computing, Vienna, Austria. URL https://www.R-project.org/.). Spearman correlations between potential predictive variables and several outcomes were determined and the matrix of correlation coefficients were plotted using the corrplot package (Taiyun Wei and Viliam Simko (2021). R package ‘corrplot’: Visualization of a Correlation Matrix (Version 0.92). Available from https://github.com/taiyun/corrplot). Correlation coefficients were reported as r. Only correlations with at least 5 measured value pairs were shown in the plots. No adjustment of p-values for multiple testing was done to avoid an inflation of type II errors. Selected single correlations were plotted using the default plot-function in R, including results for Pearson and Spearman correlations. CD4^+^ T cell activation was plotted as stimulation index (StimIndex), i.e. ratio of % of CD40L^+^4-1BB^+^ CD4^+^ T cells in stimulated samples and % of CD40L^+^4-1BB^+^ CD4^+^ T cells in unstimulated controls.

## Results

To determine the individual effects of DMTs commonly used to treat MS patients on humoral and cellular immune responses to mRNA SARS-CoV-2 vaccination, we analysed 86 DMT-treated MS patients and compared them to 17 untreated MS patients (see **Table 1** for details of DMT and clinical characteristics). Clinical histories and samples were taken prior to primary vaccination as well as one month (1m), three months (3m) and six months (6m) post primary vaccination (see Methods for details). For clinical safety reports see Suppl. Fig. 1.

**Table 1:** Clinical cohort characteristics.

| | n | female sex n [%] | mean age in years [range] | mean disease duration since manifestation in years ± SD [range] | mean treatment duration in months ± SD [range] |
| --- | --- | --- | --- | --- | --- |
| **Untreated** | 17 | 9 [53 %] | 54 [28-68] | 19±9 [3-36] | NA |
| **aCD20-BCD** | 27 | 17 [63 %] | 43 [28-58] | 14±9 [1-33] | 26±12 [2-51] |
| **Fingolimod** | 21 | 16 [76 %] | 47 [21-68] | 18±7 [7-33] | 84±53 [9-167] |
| **IFNβ** | 7 | 7 [100 %] | 42 [34-58] | 11±6 [3-20] | 102±44 [20-180] |
| **DMF** | 10 | 8 [80 %] | 45 [33-55] | 15±9 [1-29] | 90±28 [12-155] |
| **GA** | 8 | 6 [75 %] | 47 [34-61] | 13±8 [1-26] | 99±32 [15-144] |
| **TFN** | 9 | 7 [78 %] | 46 [31-55] | 11±7 [1-20] | 81±50 [14-188] |
| **NTZ** | 4 | 2 [50 %] | 45 [31-56] | 13±12 [2-26] | 55±21 [13-70] |

Interferon- $\beta$ (IFNβ ); dimethyl fumarate (DMF); glatiramer acetate (GA); teriflunomide (TFN); natalizumab (NTZ); anti-CD20-based B cell depletion (aCD20-BCD); number (n); standard deviation (SD); not applicable (NA).

### Impaired seroconversion after mRNA vaccination in aCD20-BCD- and fingolimod-treated patients

Compared with untreated MS patients, MS patients receiving aCD20-BCD therapies or fingolimod showed significantly lower or no anti-S1 IgG levels at 1m and 3m post primary vaccination (**Fig. 2a**). Ten of 11 (91%) untreated MS patients seroconverted after primary vaccination, the 11^th^ patient after secondary vaccination. In patients receiving DMTs, except for aCD20-BCD and fingolimod, seroconversion was comparable to untreated patients with conversion rates ranging from 67% to 100% after the primary vaccination, depending on the treatment group, and a 100% seroconversion rate after the secondary vaccination. In contrast to untreated MS patients, MS patients on aCD20-BCD therapies and fingolimod demonstrated significantly reduced seroconversion rates. In detail, 2 of 24 (8%) aCD20-BCD-treated patients seroconverted at 1m (p = 0.000004) and 5 of 21 (25%) at 3m (p = 0.00007); 0 of 17 (0%) fingolimod-treated patients seroconverted at 1m (p = 0.0000008) and 7 of 21 (33%) at 3m (p = 0.0004) post primary vaccination. Of note, anti-S1 IgG of seroconverted fingolimod-treated patients remained on a lower level compared to all other patients. Next, we used SARS-CoV-2 whole spike peptide arrays to determine whether antibodies to spike epitopes other than those in the spike glycoprotein S1 subunit were induced in MS patients treated with aCD20-BCD or fingolimod. Compared to untreated MS patients, these patients showed a trend toward a less diverse response pattern to the spike glycoprotein S2 subunit (**Fig. 2b**). However, both aCD20-BCD- and fingolimod-treated patients who lacked positive anti-S1 IgG levels were also devoid of antibody binding to other spike peptide fragments (**Fig. 2c**).

**Fig. 2:**
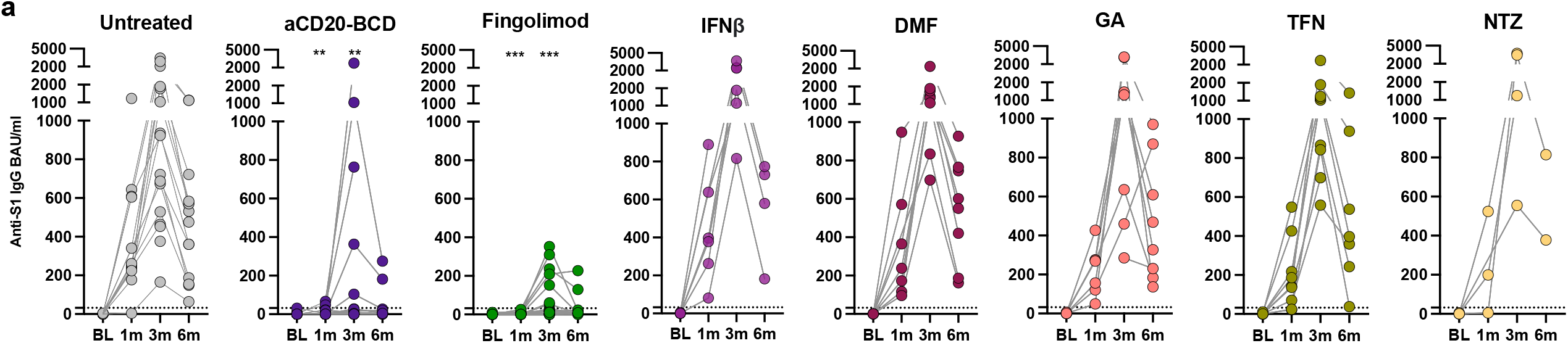
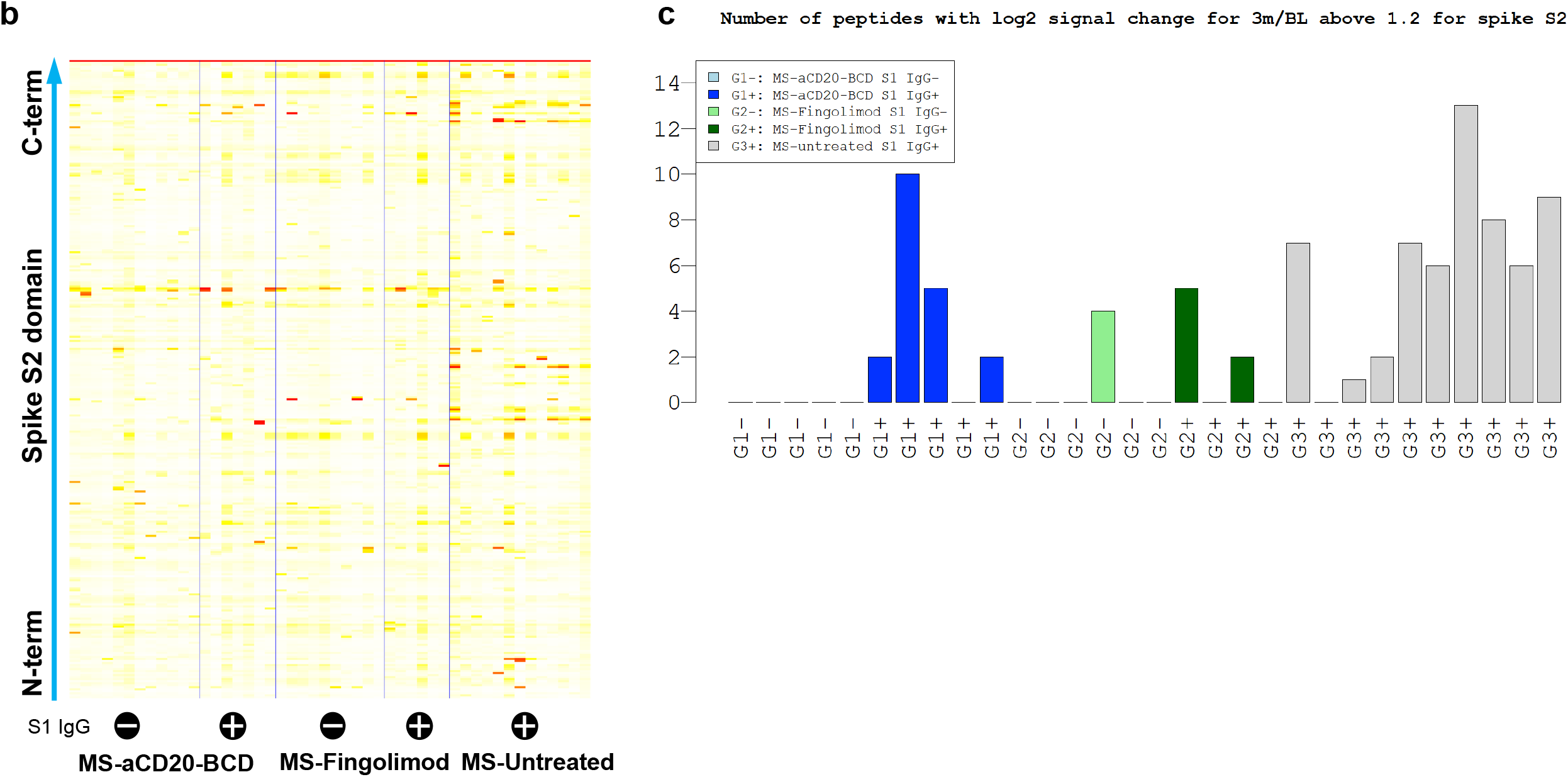
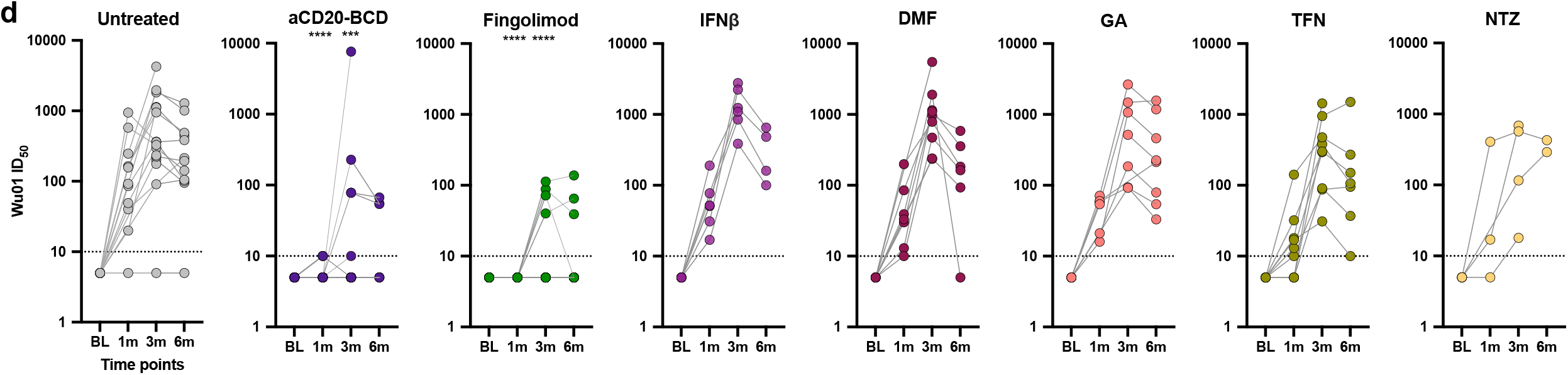
Decreased humoral responses after SARS-CoV-2 mRNA vaccination in aCD20-BCD- and fingolimod-treated patients. **a and d**, anti-S1 IgG response **(a)** and Wu01 spike neutralizing capacity **(d)** at baseline (BL) and 1m, 3m and 6m post primary vaccination per treatment group. Positivity thresholds: >31 Binding Antibody Units (BAU)/ml for anti-S1 IgG. > 10 ID_50_ for Wu01 spike neutralization. Serum ID_50_ values less than the lowest serum dilution tested (1:10) were assigned a value of 5 for plotting the graph and for statistical analysis. Kruskal-Wallis test followed by Dunn’s multiple comparisons test were performed to test treatment groups in comparison to untreated patients at the respective time points. Significance was reported as: *p ≤ 0.05, **p ≤ 0.01, ***p ≤ 0.001, ****p ≤ 0.0001. Non-significant results were not reported. **c**, peptide array results from SARS-CoV-2 S2 subunit peptides from sera from sub-cohorts of the indicated MS treatment groups (MS-aCD20-BCD n=8, MS-fingolimod n=10, MS-untreated n=9), respectively, with (+) or without (-) anti-S1 IgG antibodies. **d**, number of peptides with log2-fold signal change for 3m/BL >1.2 for all S2 peptides depicted according to sub-cohorts of the indicated treatment group (MS-aCD20-BCD n=8, MS-fingolimod n=10, MS-untreated n=9) with (+) or without (-) anti-S1 IgG antibodies.

To further investigate humoral features of the mRNA vaccine-induced immune response, we examined serum neutralisation capacities using a pseudovirus neutralization assay with the Wu01 spike protein, which is the vaccine strain (**Fig. 2d**). Overall, anti-S1 IgG levels correlated positively with the serum ability to neutralize Wu01 spike at 3m (with a spearman r value of 0.87 for untreated MS patients, 0.68 for IFNβ, 0.64 for DMF, 0.48 for GA, 0.46 for TFN, −0.11 for NTZ, 0.2 for aCD20-BCD, and 0.64 for fingolimod; **Fig. 2a and d**; Suppl. Fig. 2). The untreated MS cohort showed the highest serum neutralizing capacity with a geometric mean serum ID_50_ titre of 382 at 3m post primary vaccination. IFNβ, DMF, GA, TFN and NTZ treatment did not affect development of serum neutralizing activity at 3m. In contrast, treatment with aCD20-BCD therapies and fingolimod reduced the geometric mean serum neutralizing titres at 3m by 35- and 48-fold, respectively, compared to untreated MS patients. In stark contrast to all other groups and consistent with anti-S1 IgG production, none of the aCD20-BCD- or fingolimod-treated patients showed neutralizing serum capacity already after the first vaccination. Notably, while 7 of 21 (33%) fingolimod-treated patients mounted positive anti-S1 IgG levels after secondary vaccination (at 3m), only 4 of 21 (19 %) showed any detectable neutralizing activity with an ID_50_ at this 3m time point. No such discrepancy was found for aCD20-BCD. These findings recapitulate previous findings that antibody responses to SARS-CoV-2 mRNA vaccine are attenuated in patients receiving aCD20-BCD therapy. Importantly, they expand our knowledge of fingolimod-treated patients by showing that their antibody responses are virtually absent and, if present, lack maturation reflected by the lack of neutralizing activity.

### Impaired B cell responses after SARS-CoV-2 mRNA vaccination in aCD20-BCD- and fingolimod-treated MS patients

To define the magnitude of the receptor-binding domain (RBD)- and S2-specific B cell response in our MS patient cohorts post vaccination, we used direct staining of B cells with fluorescently labelled antigens and flow cytometry. Consistent with the serologic response, only a fraction of patients on aCD20-BCD therapies and fingolimod exhibited circulating RBD- (aCD20-BCD 7%, fingolimod 50%) and S2- (aCD20-BCD 14%, fingolimod 50%) specific B cells after the secondary vaccination (at 3m) and only in low numbers (**Fig. 3a and b**). For the other treatment groups, no significant difference in RBD- and S2-specific B cell responses was observed compared to untreated MS patients. B cell compartmentalization was similar in untreated patients and patients on DMTs except, again, for aCD20-BCD therapies and fingolimod. Most antigen-specific B cells were memory B cells regardless of S2- or RBD-specificity but the memory compartment was larger for S2-specific B cells (**Fig. 3c**). The S2 subunit of the SARS-CoV-2 spike glycoprotein is more homologous to other human coronaviruses than the S1 subunit suggesting cross-reactive memory B cell engagement as a cause of this observation^21,22,23,21,24^. For aCD20-BCD-treated MS patients, none of the few RBD-specific B cells (detectable in only 2 patients) were memory B cells. For fingolimod-treated patients, we observed a shift towards a naïve B cell phenotype of both RBD- and S2-specific B cells (detectable in only 8 patients) compared to the other MS patients. Overall, these data demonstrate and confirm that humoral responses are impaired in patients treated with aCD20-BCD. Unexpectedly, and extending previous observations, humoral and B-cell responses in MS patients treated with fingolimod were almost completely absent or, if present, at lower levels and biased in their composition.

**Fig. 3:**
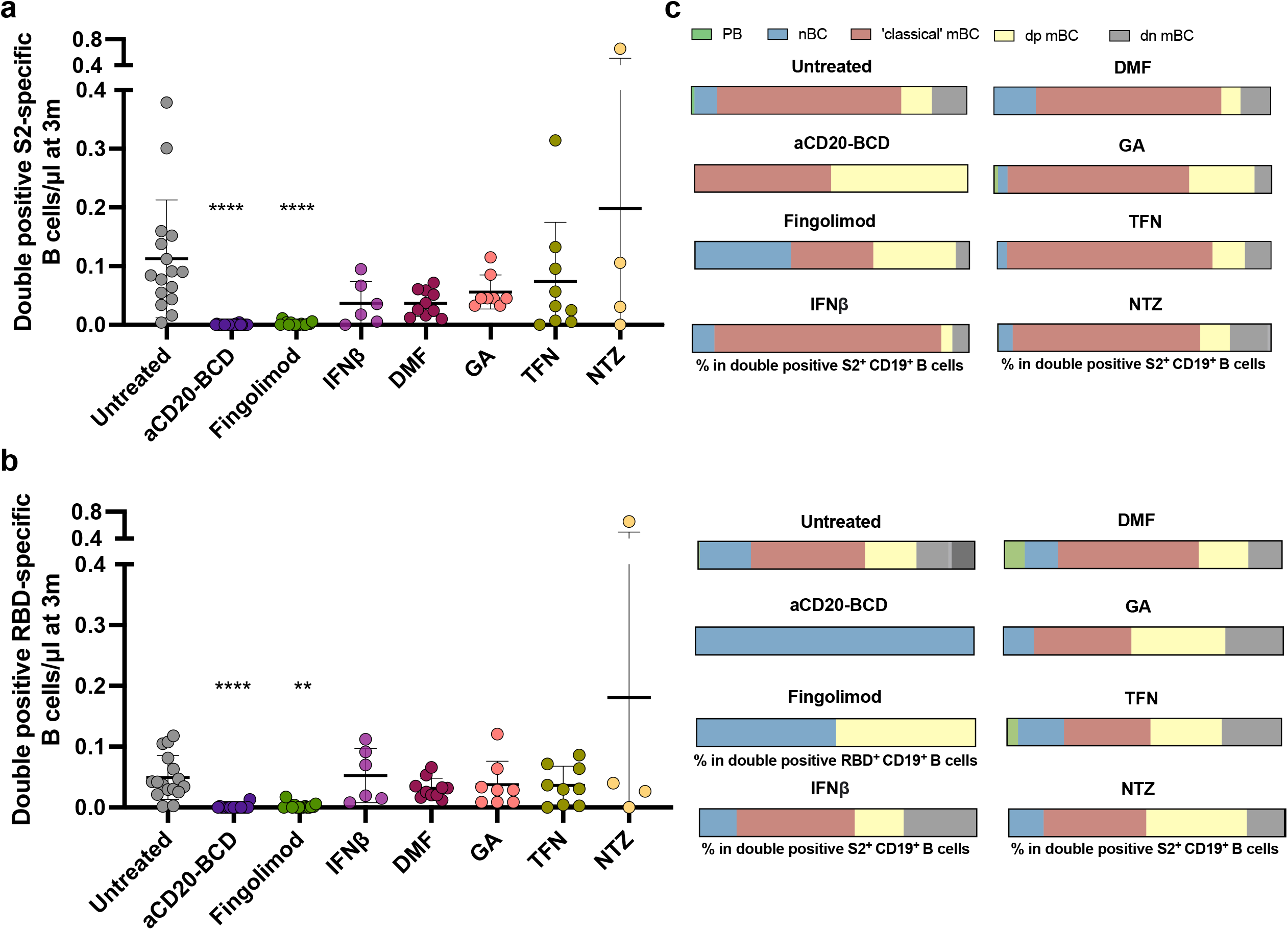
Decreased and naïve B cell-shifted RBD- and S2-specific B cell responses in aCD20-BCD- and fingolimod-treated MS patients. **a and b**, absolute numbers of RBD-specific **(a)** and S2-specific **(b)** CD19^+^ B cells at 3m post primary vaccination for each treatment group. Mean and standard deviation are indicated. Kruskal-Wallis test followed by Dunn’s multiple comparisons test performed to test treatment groups in comparison to untreated patients at the respective time points. Significance was reported as: *p ≤ 0.05, **p ≤ 0.01, ***p ≤ 0.001, ****p ≤ 0.0001. Non-significant results were not reported. **c**, frequencies of CD19^+^CD20^low/-^CD27^++^CD38^++^ plasma blasts (PB), CD19^+^CD20^+^CD27^-^ IgD^+^ naïve B cells (nBC), CD19^+^CD20^+^CD27^+^ IgD^-^ memory B cells (mBC), CD19^+^CD20^+^CD27^+^ IgD^+^ mBC (dp mBC) and CD19^+^CD20^+^CD27^-^ IgD^-^ B cells (dn mBC) in RBD-specific and S2-specific CD19^+^ B cells per treatment group. B cell gating is shown in Suppl. Fig. 4.

### Lack of vaccine-induced spike-reactive CD4^+^ T cell responses in fingolimod-treated MS patients

Robust T cell responses are associated with improved survival in COVID-19 patients with hematologic malignancies including patients receiving BCD therapies^25,26^. Given the poor humoral responses in MS patients treated with aCD20-BCD and fingolimod, we next examined CD4^+^ T cell S-I and S-II reactivity following primary and secondary SARS-CoV-2 mRNA vaccination (**Fig. 4a and b**). All MS patients without treatment and on DMTs except fingolimod demonstrated a strong increase in S-I- and S-II-reactive CD4^+^ T cells after primary vaccination. In contrast, fingolimod-treated patients showed significantly reduced or no S-I- and S-II-reactive T cell responses. Only 1 of 17 (6%) fingolimod-treated patients had a weak CD4^+^ T cell response to the S-I peptide pool after the primary vaccination and 3 of 17 (18%) to S-II. The secondary vaccination triggered low responses in 3 of 21 (14%) fingolimod-treated patients to S-I and S-II. As fingolimod treatment affects the composition of circulating T cell subsets^27,28^, we assumed an impact on the detected vaccination responses. Therefore, we next examined the effect of fingolimod on the T cell populations of our cohorts. As expected, fingolimod-treated patients showed significantly fewer circulating CD3^+^ T cells and a CD4/CD8 subset shift in favour of CD8^+^ T cells (**Fig. 4c and d**). In addition, we observed significantly more signalling lymphocyte activation molecule family member 7 positive (SLAMF7^+^) cytotoxic CD4^+^ and CD8^+^ T cells^29^ in these patients (**Fig. 4e**) indicating a dominance of cytotoxic T cells and very few circulating CD4^+^ helper T cells. The distribution of circulating T cell subsets before vaccination remained largely unchanged post vaccination in all groups although the absolute number of CD3^+^ T cells varied (**Fig. 4c and d**). Overall, spike-reactive T cell responses were severely impaired in MS patients treated with fingolimod, which is in stark contrast to untreated patients and patients on other DMTs including aCD20-BCD. In patients with an impaired humoral response, it has been discussed that the spike-reactive T cells are particularly relevant for protection ^25,26^. Our data suggest that fingolimod-treated MS patients appear to lack this safety net.

**Fig. 4:**
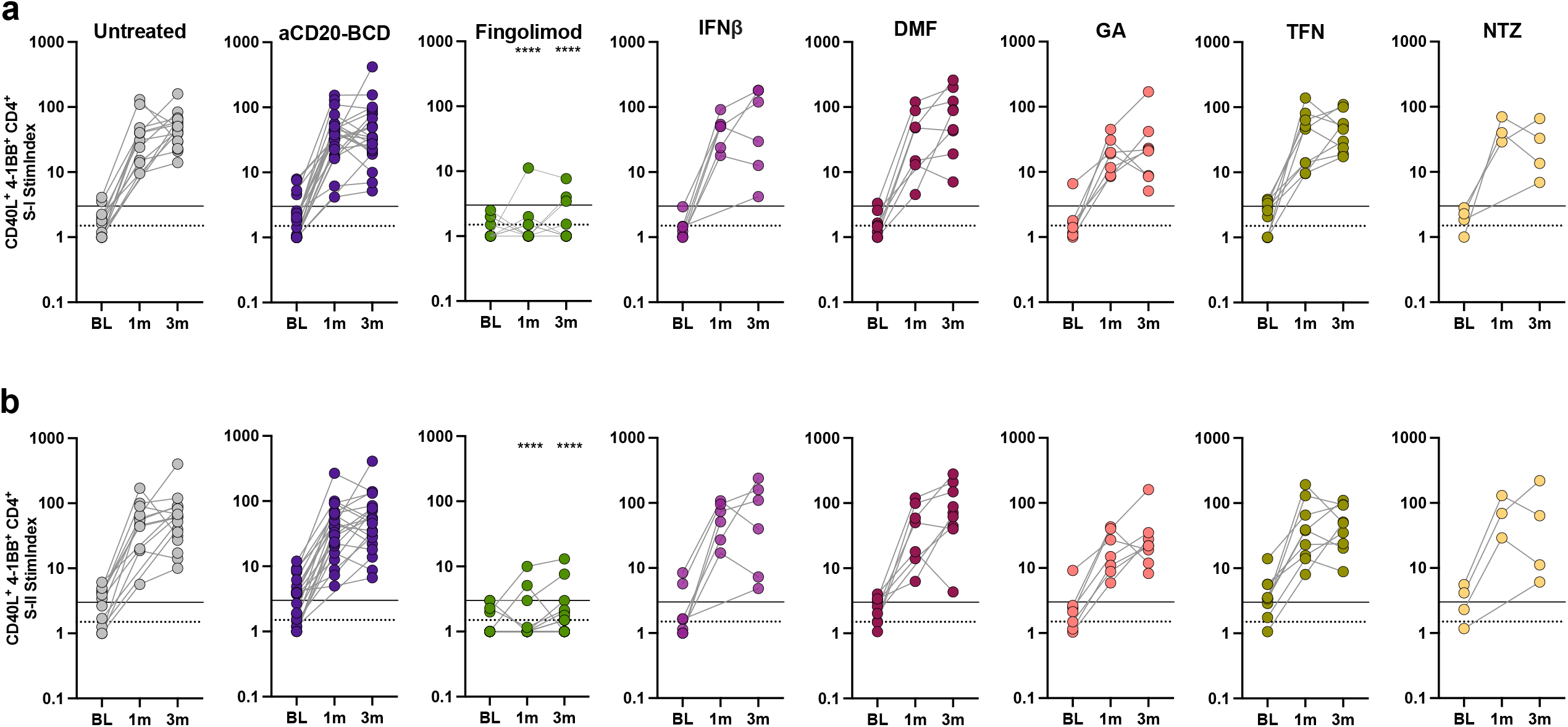
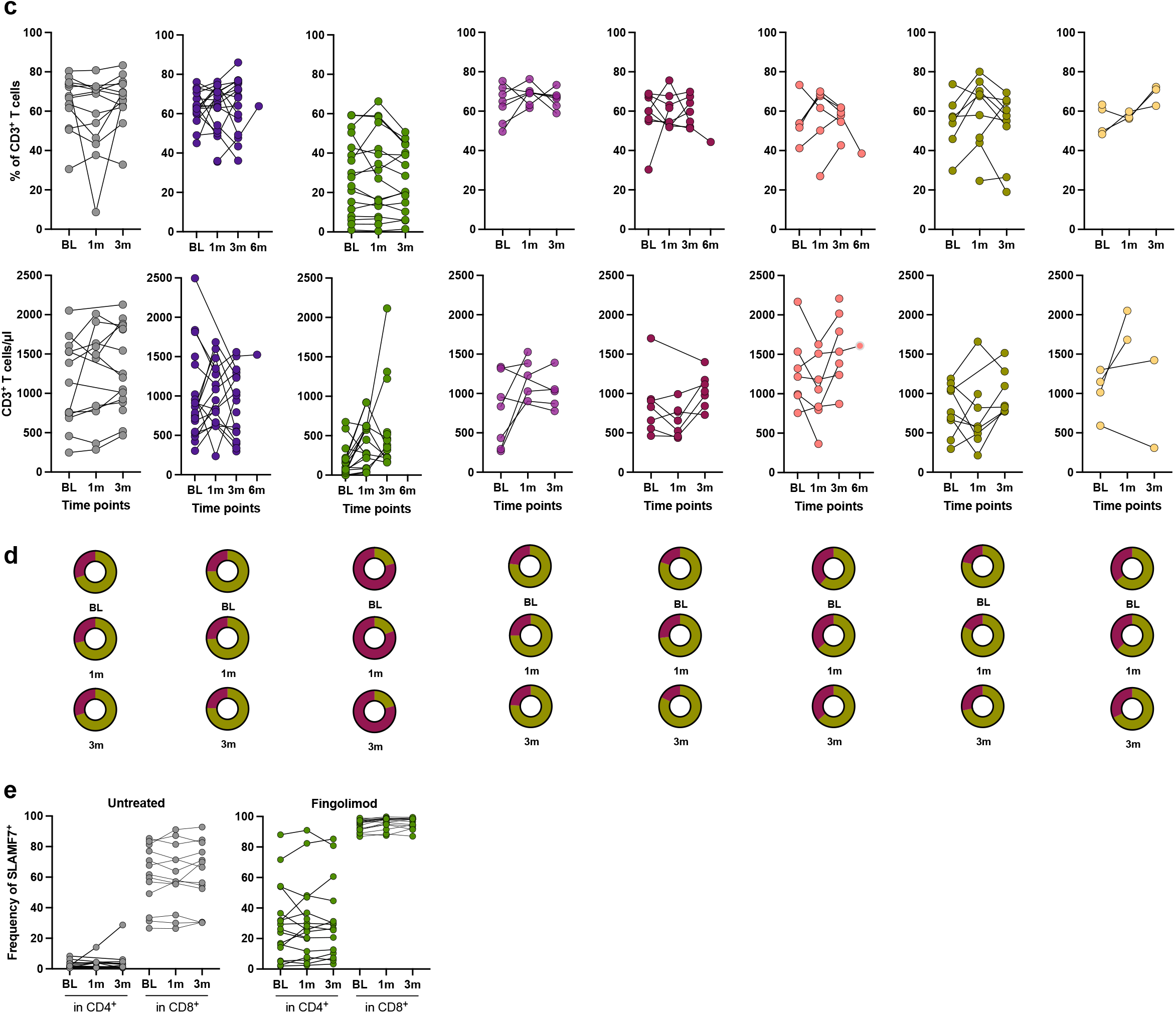
Impaired S-I- and S-II-reactive CD4 T cell responses in fingolimod-treated MS patients. **a and b**, S-I-specific **(a)** and S-II-specific **(b)** CD4^+^ T cell reactivity at BL, 1m and 3m post first vaccination per treatment group. Lines indicate stimulation indices (StimIndex) of 1.5 (positive with uncertainty; dotted line) and 3.0 (definitely positive; solid line). T cell gating is shown in Suppl. Fig. 5. Kruskal-Wallis test followed by Dunn’s multiple comparisons test performed to test treatment groups in comparison to untreated patients at the respective time points. Significance was reported as: *p ≤ 0.05, **p ≤ 0.01, ***p ≤ 0.001, ****p ≤ 0.0001. Non-significant results were not reported. **c**, frequencies and absolute CD3^+^ T cell number at BL, 1m and 3m after primary vaccination per treatment group. **d**, frequencies of CD4^+^ (violet) and CD8^+^ (green) cells in CD3^+^ T cells at BL, 1m and 3m post first vaccination per treatment group. **e**, frequencies of SLAMF7^+^ cells in CD4^+^ or CD8^+^ T cells, respectively, at BL, 1m and 3m for patients treated with fingolimod compared to untreated patients.

### Humoral and cellular immune response coordination in fingolimod-treated MS patients

Next, we performed correlation analyses to gain insight into which factors influencing the individual immunologic health status may be associated with the partial or complete absence of vaccine-induced immune responses in aCD20-BCD- and fingolimod-treated MS patients. These analyses revealed negative correlation trends between antibody induction and disease duration, and positive correlation trends between antibody induction and the amounts of B cells and T cells before vaccination (**Fig. 5a-c**). As shown previously, we found that anti-S1 IgG levels post vaccination in patients receiving aCD20-BCD therapies showed a significant positive correlation with the frequency and absolute number of B cells before vaccination (**Fig. 5b**, Suppl. Fig. 3). In fingolimod-treated patients, B and T cell counts at baseline showed a significant positive correlation with the presence of S2-specific B cells at 1m post primary vaccination but, importantly, not with anti-S1 IgG induction or neutralizing capacity. Instead, we found a strong negative correlation between treatment duration and (neutralizing) antibody induction in these patients (**Fig. 5c and d**) with a cut-off for IgG antibody induction at 29 months.

**Fig. 5:**
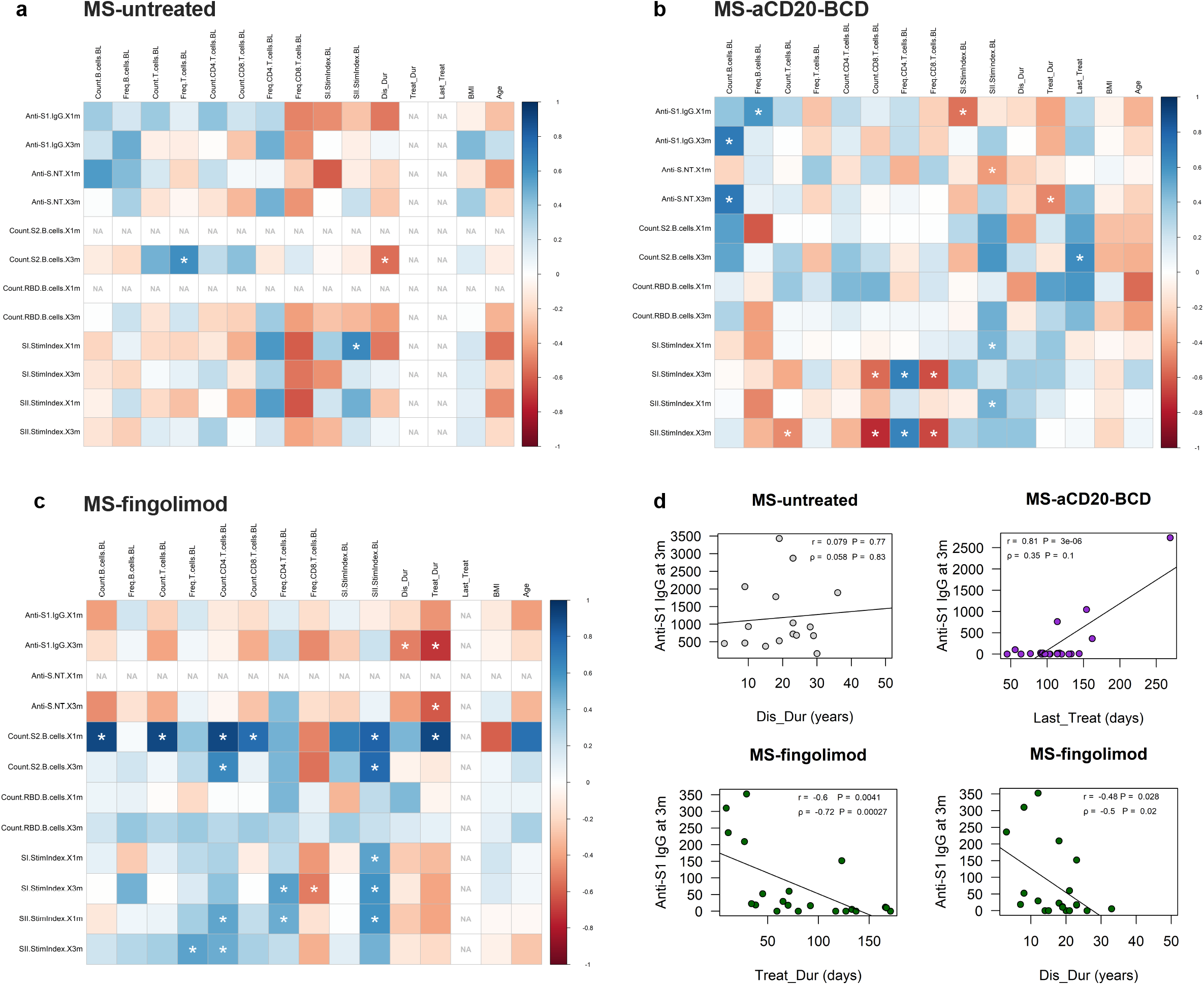
SARS-CoV-2 mRNA vaccination outcome to immune constitution correlation. **a - c**, multiple correlation analyses for untreated MS patients **(a)**, MS-aCD20-BCD patients **(b)**, and MS-fingolimod **(c)**. The y-axis represents vaccination outcome variables, the x-axis potential correlation variables. The colour of the result depicts the magnitude and direction of the respective correlation as indicated in the key on the right side. Spearman correlations with a p-value below 0.05 were considered significant and indicated by white asterisks. Correlations with less than five measured value pairs were excluded due to low robustness and indicated by “NA”. **d**, simple linear correlation for anti-S1-IgG at 3m and disease duration (Dis_Dur; MS-untreated), days since last treatment administration (Last_Treat; MS-aCD20-BCD), disease duration and overall treatment duration (Treat_Dur; MS-fingolimod; from upper left clock-wise).

## Discussion

We here provide a detailed analysis of primary and secondary immune responses to SARS-CoV-2 mRNA vaccination in MS patients treated with various immunomodulatory treatment regimens including the S1P receptor functional antagonist fingolimod and aCD20-BCD therapies. SARS-CoV-2 vaccines mitigate symptom severity of COVID-19 by inducing specific T cell and B cell memory^30,31^ but how B and T cells individually act to prevent severe COVID-19 is so far unclear. The higher incidence of severe COVID-19 disease in aCD20-BCD-treated MS patients suggests that a lack of specific antibodies and B cells can severely impair COVID-19 immunity^32,6^. However, antibody responses to SARS-CoV-2 appear to be short-lived even in healthy individuals without known immunodeficiency, thus shifting the focus to memory B and T cellular immune memory to provide long-term protection from severe disease. T cell responses to SARS-CoV-2 vaccination have been shown to be stable and to play a central role in protection, enhancement of the antibody response, and viral clearance^20,33^. Consistently, reduced T cell responses against SARS-CoV-2 have also been associated with severe disease^34^. In patients undergoing aCD20-BCD therapy, who consequently lack B cells and are unable to mount specific humoral immune responses, T cell responses are promising indicators of protective immune responses by providing B cell-independent protection from severe COVID-19.

Our findings demonstrate that untreated MS patients and patients on DMT, except for aCD20-BCD therapies and fingolimod, mounted humoral immune responses to SARS-CoV-2 vaccination on a comparable level to that of normal healthy donors^13,20,35^. Interestingly, the neutralizing antibody capacity in some aCD20-BCD-treated patients remained at a low level even after the second vaccination despite the induction of average levels of anti-S1 IgG suggesting impaired affinity maturation, in line with selective defects in antigen-specific circulating follicular helper T cells in this treatment group^13^ or premature termination of the immune response. Our results from patients on aCD20-BCD therapy are in accordance with previous reports showing that anti-S1 IgG levels correlated with the absolute B cell number in the peripheral blood and also, but to a lesser extent, with duration since the last aCD20-BCD treatment administration prior to vaccination, and finally that aCD20-BCD has no effect on the principal ability to elicit SARS-CoV-2 spike-specific T-cell responses^13, 16, 36^.

In contrast to all other treatment groups, the majority of fingolimod-treated MS patients developed neither humoral nor CD4^+^ T cellular immune responses to SARS-CoV-2 vaccination. In line with low or no humoral responses, fingolimod-treated patients demonstrated absent or drastically reduced numbers of S2- and RBD-specific B cells. Interestingly, fingolimod-treated patients did not report significantly fewer systemic side effects than individuals with measurable humoral and cellular immune responses. Fingolimod is a compound molecule which sequesters lymphocytes in secondary lymphoid organs (SLO) and inflammatory tissues and thus hinders lymphocyte recirculation into the periphery. It also facilitates an increased T cell responsiveness to ligands of the CCR7 receptor^37^. T cells utilize CCR7 to home and migrate within the T cell zone and B cells express that receptor to position at the T:B border. To successfully mount a functional immune response, T and B cells must migrate from niche to niche and zone to zone in a rapid and highly regulated manner^38^, a process that is abolished by fingolimod. Earlier studies also demonstrate effects of fingolimod on the mobility of dendritic cells^39^, which taken all together may explain the lack of immune response in these patients and underscores the importance of immune cell migration in the coordinated nature of functional immune responses.

Interestingly, observations in mice showed that fingolimod leads to sequestration of T cells in SLO but not that fingolimod completely abrogates specific immune responses^40^. However, in these experiments mice were treated with fingolimod only over a short period of time. Few studies have examined immune responses in humans during treatment with fingolimod but the absence of humoral immune responses to SARS-CoV-2 vaccination has been reported^16^. In contrast to our findings here, a recently published study did not show a clear lack of immune response^41^. In this study, an ELISPOT IFN*γ* assay was used to determine the amount of antigen-specific T cells. ELISPOT IFN*γ* assays can be less sensitive for antigen-specific T cells as these assays are unable to distinguish between antigen-specific and bystander IFN*γ*-secreting cells which may be present at increased levels in fingolimod-treated patients due to the shift toward cytotoxic effector (SLAMF7^+^) T cells. The activation-induced marker (AIM)-based assay used here assesses the entirety of S-I- and S-II-reactive CD4^+^ T cells based on their TCR engagement and independent of the cytokines they secrete. A notable limitation, however, for both assays is the skewing of peripheral T cell subsets toward CD8^+^ T cells in fingolimod-treated patients. As a result, for the same number of cells seeded for stimulation, fewer CD4^+^ T helper cells are seeded than in all other patient groups, thereby affecting detection limits. Further studies will be required to clarify the discrepancies and guide precautionary measures for these patients.

While in aCD20-BCD-treated patients, vaccination efficacy correlated with the time elapsed since last treatment administration and the absolute number of B cells at the time of vaccination, in fingolimod-treated patients, vaccine-induced humoral responses were negatively correlated with overall treatment duration. In MS, treatment suspension is often not feasible as reduced immunomodulation increases the risk of autoimmune flare-ups. Thus, there are currently very few options to improve the pre-conditions for successful vaccination outcomes in these patients, which poses a high risk to them. It remains unclear why a few fingolimod-treated patients mounted (low) humoral and cellular immune responses after secondary vaccination while most others did not.

In summary, in striking contrast to all other DMTs, the majority of patients treated with fingolimod appear to be unable to mount specific B and T cell responses to SARS-CoV-2 vaccination. Accordingly, fingolimod-treated patients may require frequent immune check-ups and further means of protection, which becomes increasingly relevant for clinical practice due to the recent approval of three new S1P receptor antagonists (siponimod, ozanimod, ponesimod) with broad applications in different types of MS and thus a growing number of patients.

## Data Availability

The data that support the findings of this study are available from the corresponding author upon reasonable request.

## Acknowledgments

We thank the CCC Study Group E. Baysal, T. Panne, F. Legler, M. Girod, B. Bohnen, T. Nguyen, J. Schmitz, R. Klemencic, L. Hintze, N. Avinc, P. Resch, A. Maraj, A. Farghaly, P. Schulz, N. Matuschewski, K. Gutwenger, I. Katsianas and C. Dedieu for their contributions to donor recruitment, sample processing and measurement. Furthermore, we thank A. Hetey for support with maintaining the REDCap database and N. Brindle for critical reading of the manuscript.

## Authors’ contribution

Conceptualization: LMA, FP, JBS, AT, CGT. Data Analysis: LMA, JB, FF, KV, KJ, MM, AM, AB, PH, UR, CGS. Funding acquisition: AT. Resources: LMA, LL, LH, MD, BK, CB, AN, KDLR, CR, BS, LES, FK. Writing: LMA, FP, JBS, AT, CGT. LMA and CGT have verified the manuscript’s data.

## Declaration of interests

PH and UR are employed by JPT Peptide Technologies. All other authors declare no competing interests.

## Notes

### Funding Statement

This study was funded by the German Ministry of Health.

### Author Declarations

The ethics committee of the Charite - Universitaetsmedizin Berlin gave ethical approval for this work.

