## Supplemental Material for "SARS-CoV-2 mRNA vaccination fails to elicit humoral and cellular immune responses in multiple sclerosis patients receiving fingolimod"

### Supplemental Methods

#### Assessment of vaccine reactogenicity and severe adverse events

At follow-up visits, participants were asked to report on occurrence and severity of local and systemic reactions and severe adverse events (SAEs) during the first seven days after the respective vaccination. SAEs included worsening of MS symptoms. Reactogenicity and SAEs were assessed by a modified US Food and Drug Administration toxicity scale rating from mild (does not interfere with daily activities) to moderate (interferes with daily activities) and severe (daily activities no longer feasible)[39].

#### SARS-CoV-2 pseudovirus neutralization assay

SARS-CoV-2 pseudoviruses were generated by co-transfection of plasmids encoding HIV Tat, HIV Gag/Pol, HIV Rev, luciferase followed by an IRES and ZsGreen, and the SARS-CoV-2 spike protein (Wu01 spike, EPI_ISL_406716) into HEK 293T cells using FuGENE 6 Transfection Reagent (Promega). Virus culture supernatant was harvested at 48 h and 72 h post transfection and stored at -80°C till use. Harvested virus was titrated by infecting 293T expressing ACE2[42] and after a 48-hour incubation at 37°C and 5% CO2, luciferase activity was determined after addition of luciferin/lysis buffer (10 mM MgCl2, 0.3 mM ATP, 0.5 mM Coenzyme A, 17 mM IGEPAL (all Sigma-Aldrich), and 1 mM D-Luciferin (GoldBio) in Tris-HCL) using the Tristar microplate reader (Berthold). Neutralization assays were performed as described before. Briefly, 3-fold serial dilutions of serum (1:10 starting dilution) were co-incubated with pseudovirus supernatants for 1 h at 37°C, following which 293T-ACE-2 cells were added. After 48 h at 37°C and 5% $CO_2$, luciferase activity was determined using the luciferin/lysis buffer. Background relative light units (RLUs) of non-infected cells was subtracted and 50% inhibitory dilution ($ID_{50}$) were calculated as the serum dilution resulting in a 50% reduction in RLU compared to the untreated virus control wells. $ID_{50}$ values were calculated by plotting a non-linear fit dose response curve in GraphPad Prism 7.0.

#### SARS-CoV-2 spike epitope-specific peptide microarray

Peptides were synthesized and immobilized on peptide microarray slides as described previously. In brief, the peptides were synthesized using SPOT synthesis, cleaved from the solid support and chemoselectively immobilized on functionalized glass slides. Each peptide was deposited on the microarray in triplicates. The peptide microarrays were incubated with human sera (applied dilution 1:200) in a 96-well microarray incubation chamber for one hour at 30°C, followed by incubation with 0.1 µg/mL fluorescently labelled anti human IgG detection antibody (Jackson Immunoresearch). Washing steps were performed after each incubation step with 0.1 % Tween-20 in 1x TBS. After the final incubation step the microarrays were washed and dried. Each microarray slide was scanned using a GenePix Scanner 4300 SL50 (Molecular Devices). Signal intensities were evaluated using GenePix Pro 7.0 analysis software (Molecular Devices). For each peptide, the MMC2 value of the three triplicates was calculated. The MMC2 value was equal to the mean value of all three instances on the microarray except when the coefficient of variation (CV) – standard-deviation divided by the mean value – was larger than 0.5. In this case the mean of the two values closest to each other (MC2) was assigned to MMC2. Further data analysis and generation of the heatmaps was performed using the statistical computing and graphics software R (Version 4.1.1, www.r-project.org).

#### *Ex vivo* T cell stimulations

In short, PBMC were stimulated with PepMix™ SARS-CoV-2 spike glycoprotein pool 1 (JPT) covering the N-terminal aa residues 1-643 (abbreviated to "S-I") and with PepMix™ SARS-CoV-2 spike glycoprotein pool 2 covering the C-terminal part (aa residues 633-1273, abbreviated to "S-II") at a final concentration of 1 µg/ml per peptide, respectively. Stimulation controls were performed with equal concentrations of DMSO in PBS (unstimulated control) and 1 µg/ml per peptide of CEFX Ultra SuperStim pool (JPT) as positive control. All approaches contained 1 µg/ml purified anti-CD28 (clone CD28.2; BD Biosciences). Incubation was performed at 37˚C, 5% $CO_2$ for 16 h in the presence of 10 µg/ml brefeldin A (Sigma-Aldrich) during the last 14h. T cell stimulations were stopped by incubation in 20mM EDTA for 5 min.

**B cell and T cell flow cytometry**

For S-I- and S-II-specific T cell analysis, antibodies were used as described before: CD3-FITC (REA613, Miltenyi), CD4-VioGreen (REA623, Miltenyi), CD8-VioBlue (REA734, Miltenyi), CD38-APC (REA671, Miltenyi), HLA-DR-PerCpVio700 (REA805, Miltenyi). For B cell status analysis: CD8 Vioblue (REA734, Miltenyi), IgD BV510 (IA6-2, Biolegend), CD14 BV570 (M5E2, Biolegend), CD21 BV605 (1048, BD), CD3 FITC (REA613, Miltenyi), SLAMF7 PE (162.1, Biolegend), CD27 PE-Dazzle594 (O323, Biolegend), HLADR Percp-Vio770 (REA780, Miltenyi), CD20 PE-Vio770 (REA780, Miltenyi), CD38 APC (REA572, Miltenyi), CD4 AlexaFluor700 (RPA-T4, Biolegend) and CD19 APC-Vio770 (REA675, Miltenyi). For intracellular staining of stimulated T cells, fixation and permeabilization were performed with eBioscience™ FoxP3 fixation and PermBuffer (Invitrogen) according to the manufacturer's protocol. Intracellular staining was carried out for 30 min in the dark at room temperature with 4-1BB-PE (REA765, Miltenyi) and CD40L-PeVio770 (REA238, Miltenyi).

Peripheral blood B and T cell subsets were quantified using the following staining: 50µl of blood were stained for 20 minutes at room temperature in the presence of 1mg/ml Beriglobin (CSL Behring) and the following antibodies at their determined optimum titration: CD4-VioBlue (REA623, Miltenyi), CD3-VioGreen (REA641, Miltenyi), HLA-A2-FITC (REA517, Miltenyi), CCR7-PE (REA108, Miltenyi), CD8-PerCP (REA734, Miltenyi), CD31-PE-Vio770 (REA730, Miltenyi), CD19-APC (REA675, Miltenyi), CD16-AF700 (REA423, Miltenyi), CD45RA-APC-Vio770 (REA1047, Miltenyi). 500µl of cold Erythrocyte lysis buffer (Buffer EL, Quiagen) were added and incubated 30 minutes on ice. 400µl of cold PBS, BSA, 2mM EDTA were then added.

### Supplemental Results

#### Supplemental Fig. 1

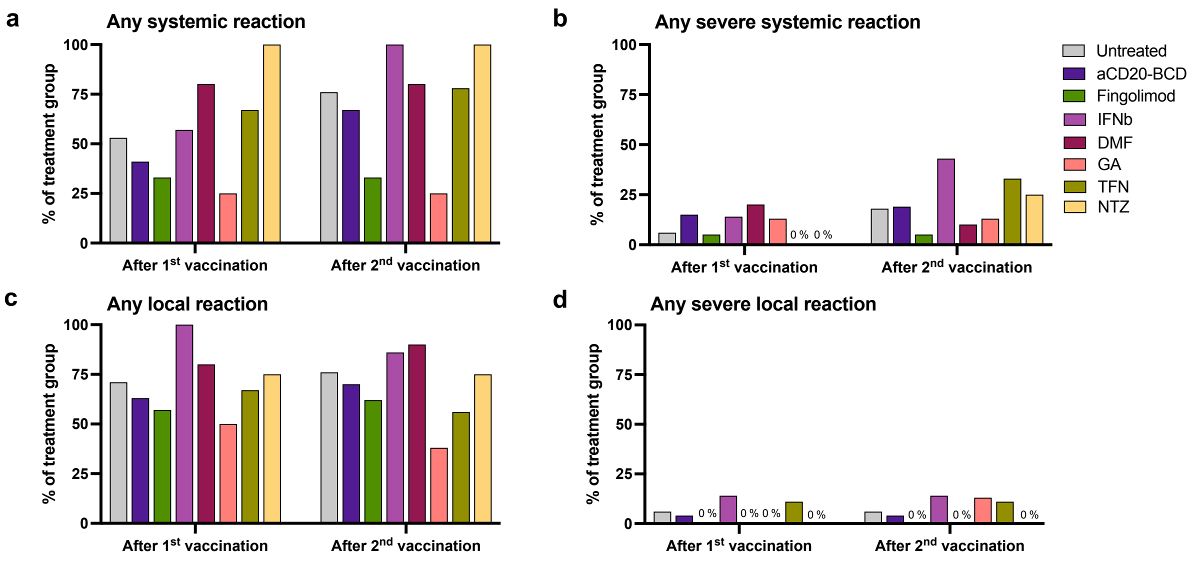


**Supplemental Fig. 1: SARS-CoV-2 mRNA vaccination-related reactogenicity. a and c,** percentage of patients grouped according to treatment with at least one systemic (**a**) or local (**c**) reaction. **b and d,** percentage of patients grouped according to treatment with at least one severe systemic (**b**) or local (**d**) reaction. 86% of all patients reported vaccination-associated side effects during the first seven days following at least one of the two vaccinations: 79% after the primary vaccination, 82% after the secondary dose, without significant differences between treatment groups. No serious adverse events were reported. Transient worsening of pre-existing neurological symptoms associated with MS was described by three patients after primary vaccination and nine patients after secondary vaccination, but no treatment group-dependent patterns were observed. Two patients on aCD20-BCD therapies tested positive for SARS-CoV-2 during the study after 1m and 3m, respectively, both with mild symptoms (WHO classification II). They were subsequently excluded from further analyses.

#### Supplemental Fig. 2

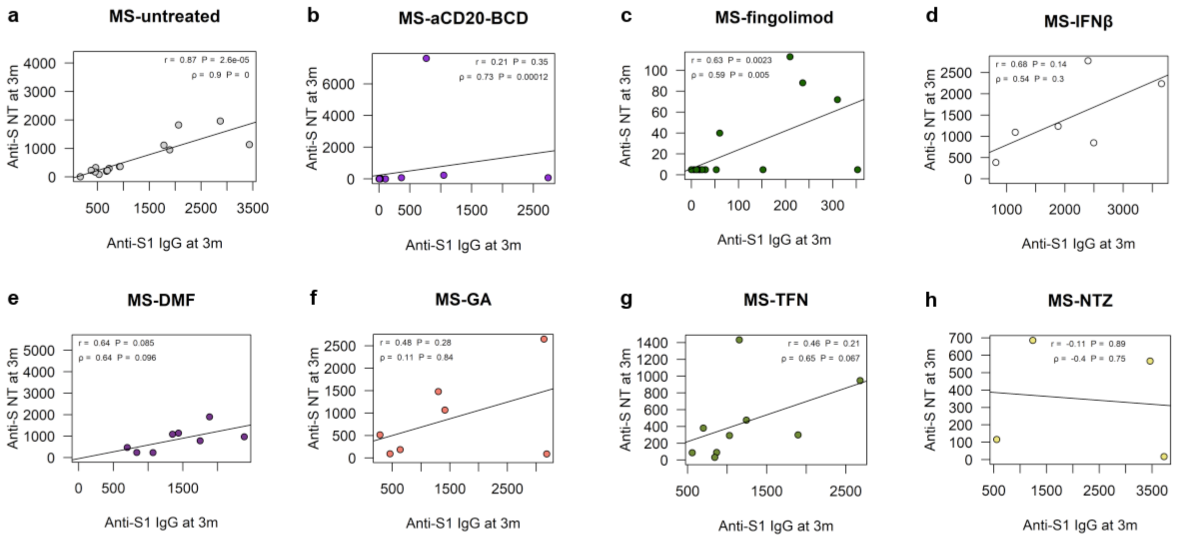

**Supplemental Fig. 2. Association of anti-S1 IgG levels and neutralizing capacity.** Correlation of anti-S1 IgG and neutralizing capacity in serum at 3m for untreated MS patients (MS-untreated) **(a)**, MS-aCD20-BCD **(b),** and MS-FTY720 **(c)**, MS-IFNβ **(d)**, MS-DMF **(e)**, MS-GA **(f)**, MS-TFN **(g)**, and MS-NTZ **(h)**. Simple linear regression tests performed per treatment; correlation coefficients depicted as lines and reported as r (for Pearson correlation) and rho (for Spearman rank correlation), respectively, with corresponding p-values.

#### Supplemental Fig. 3

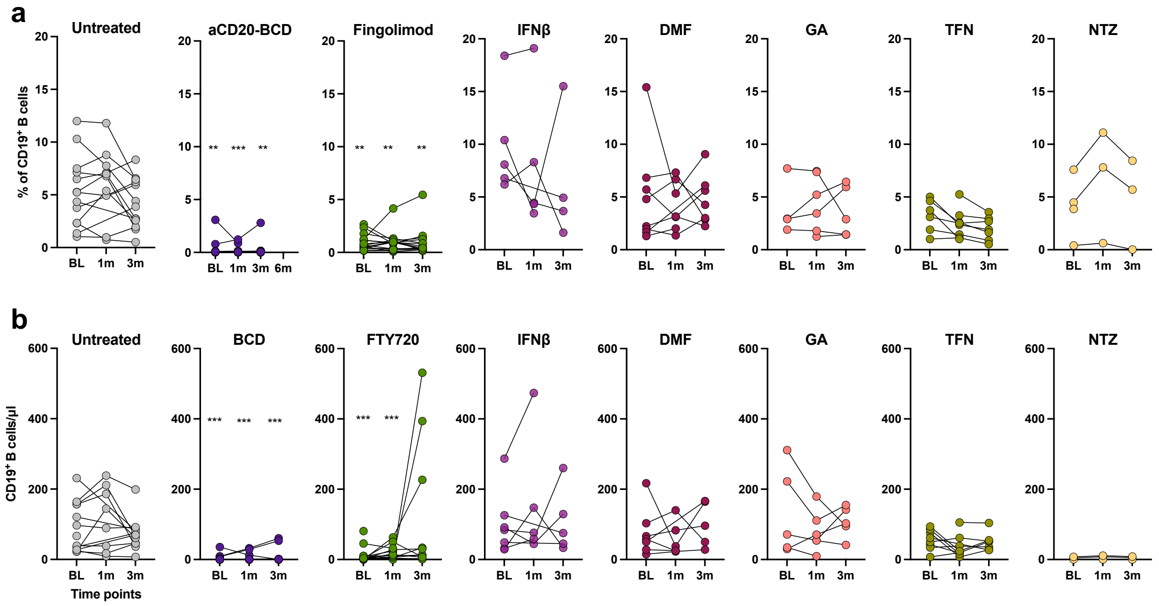


**Supplemental Fig. 3. Total number and relative frequencies of peripheral blood $CD19^+$ B cells. a)** Frequencies of $CD19^+$ B cells in total lymphocytes and **b)** total number of $CD19^+$ B cells per µl per treatment group. Kruskal-Wallis test followed by Dunn's multiple comparisons test performed to test between treatment groups at the respective time point were reported for comparison to untreated patients as: *p ≤ 0.05, **p ≤ 0.01, ***p ≤ 0.001, ****p ≤ 0.0001.

#### Supplemental Fig. 4

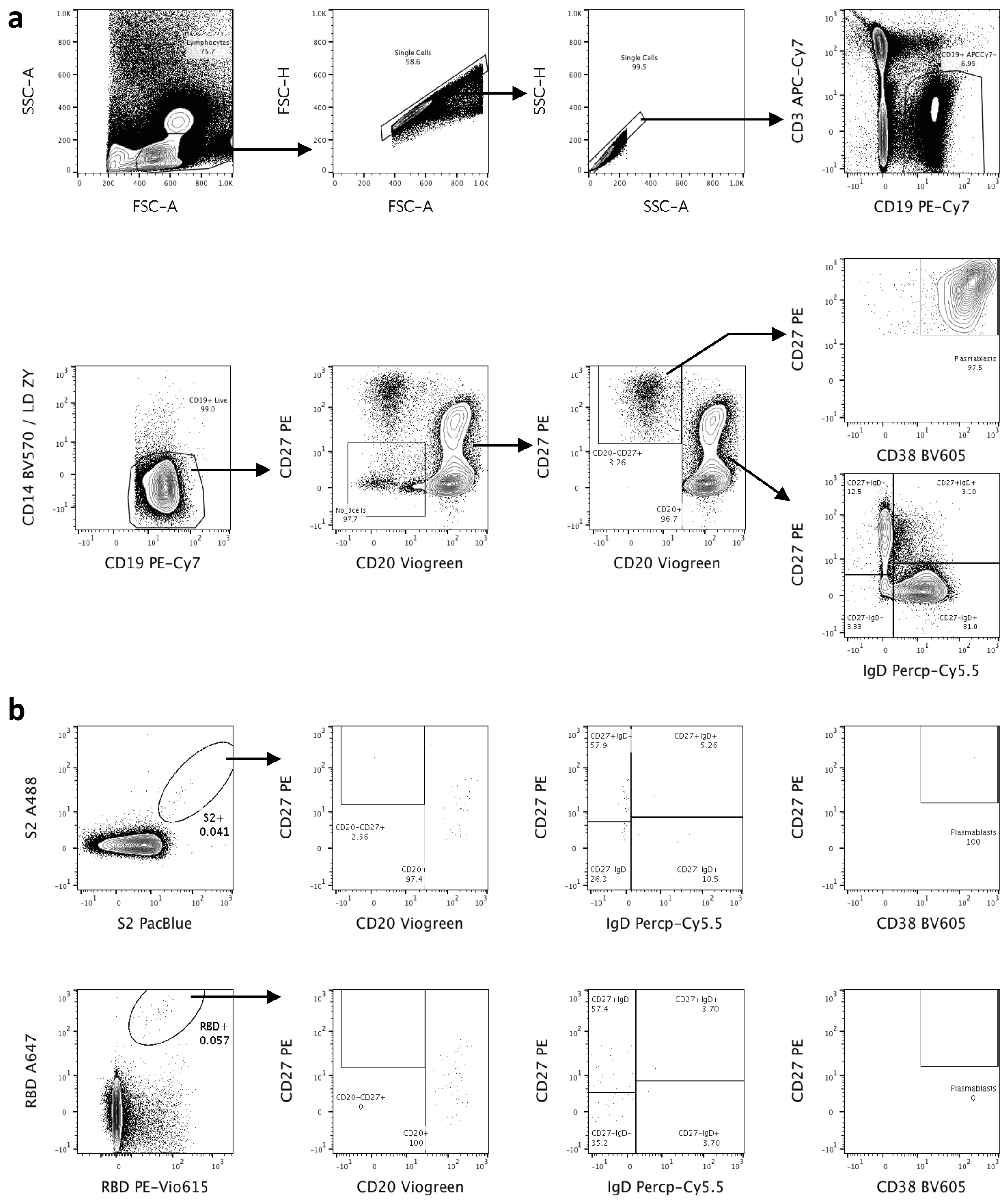


**Supplemental Fig. 4. Gating strategy of RBD- and S2-specific B cells. a,** B cells are gated from peripheral blood mononuclear cells (PBMCs; FSC-A vs SSC-A), by first excluding doublets (FSC-A vs FSC-H and SSC-A vs SSC-H), gating on $CD19^{+}CD3^{-}$ B cells, excluding both dead cells (LiveDead Zombie Yellow) and $CD14^{+}$ cells, and finally by excluding $CD20^{-}$ $CD27^{-}$ cells. Among $CD19^{+}CD3^{-}$ B cells, plasmablasts were identified as $CD20^{-/low}CD27^{++}CD38^{++}$ cells. $CD20^{+}$ B cells were further classified into subpopulations based on their CD27 and IgD expression. $CD20^{+}CD27^{+}IgD^{-}$ were classified as classical memory B cells (mBC), $CD20^{+}CD27^{+}IgD^{+}$ were classified as double positive mBC (dp mbC), $CD20^{+}CD27^{-}IgD^{-}$ were classified as double negative mBC (dn mBC), and $CD20^{+}CD27^{-}IgD^{+}$ were classified as naïve B cells (nB). **b**, exemplary contour plots showing gating of S2-specific B cells and RBD-specific B cells, and again their subsequent subpopulation classification based on their CD27 and IgD expression.

**Supplemental Fig. 5**

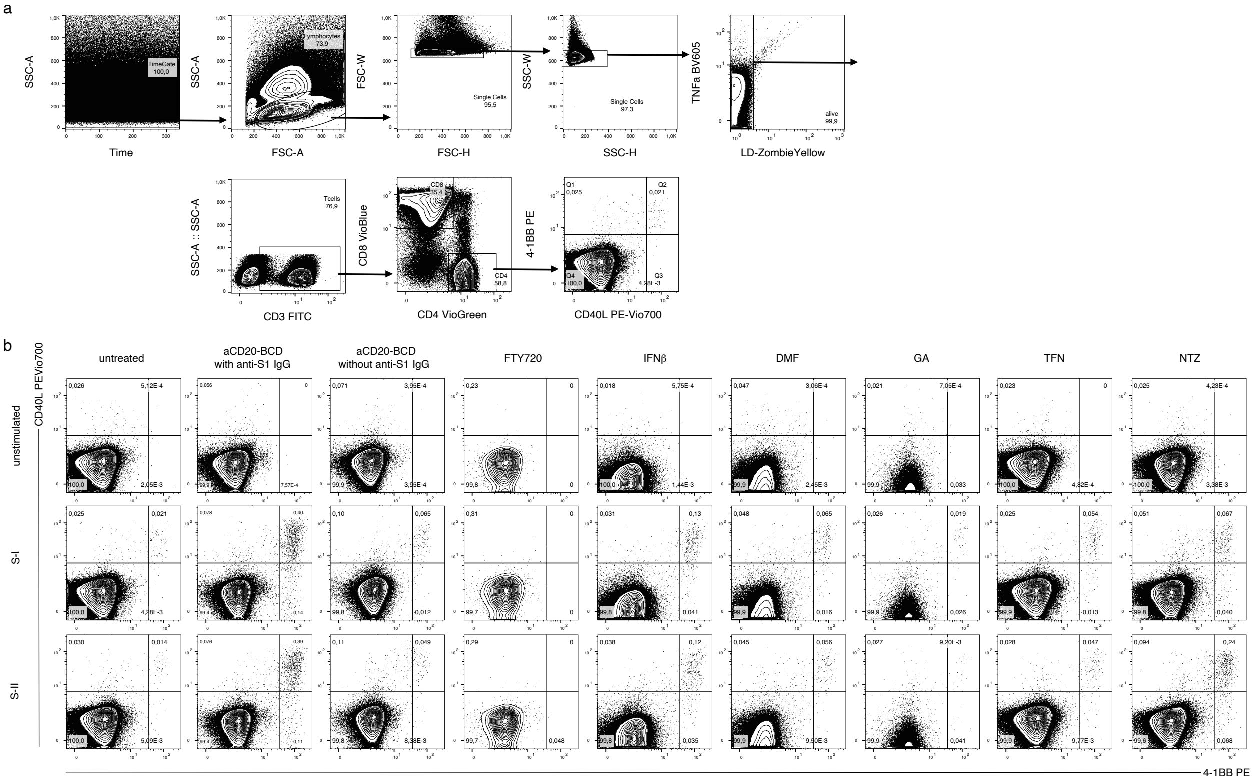


**Supplemental Fig. 5. Gating strategy of antigen-reactive CD4⁺ T cells and differences between DMT treatment groups. a**, antigen-reactive CD40L⁺4-1BB⁺CD4⁺ T cells were gated by controlling for measurement-dependent artefacts (Time vs SSC-a) from antigen-stimulated PBMC (FSC-A vs SSC-A), by first excluding doublets (FSC-H vs FSC-W and SSC-H vs SSC-W), dead cells (LiveDead Zombie Yellow), then gating on CD3⁺ T cells and CD4⁺ double positive cells. **b**, exemplary contour plots showing CD40L and 4-1BB expression in unstimulated control, S-I-, and S-II-stimulated CD4⁺ T cells, displayed for the seven different DMT treatment groups at around two months after secondary SARS-CoV-2 mRNA vaccination.

Filename: MSVX_Supplement_31012022.docx
Directory: /Users/lilmeyerarndt/Library/Containers/com.microsoft.Word/Data/Documents
Template: Normal.dotm
Title:
Subject:
Author: Meyer Arndt, Lil Antonia
Keywords:
Comments:
Creation Date: 14/01/2022 11:38:00
Change Number: 61
Last Saved On: 31/01/2022 18:33:00
Last Saved By: Meyer Arndt, Lil Antonia
Total Editing Time: 17 Minutes
Last Printed On: 31/01/2022 18:34:00
As of Last Complete Printing
Number of Pages: 7
Number of Words: 1,888
Number of Characters: 15,661 (approx.)
